# Epidemiological impact of the paediatric live attenuated influenza vaccine (LAIV) programme on group A *Streptococcus* (GAS) infections in England

**DOI:** 10.1101/2022.12.16.22283602

**Authors:** Mary A Sinnathamby, Fiona Warburton, Rebecca Guy, Nick Andrews, Theresa Lamagni, Conall Watson, Jamie Lopez Bernal

## Abstract

Influenza is known to predispose to secondary bacterial infections including group A streptococcal infection (GAS) and invasive (iGAS) disease.

The universal paediatric live attenuated influenza vaccine (LAIV) programme was introduced in England during the 2013/14 influenza season to directly protect children as well as indirectly protect the wider population through reduction in transmission. Nationally, the programme was implemented incrementally introducing cohorts of children from pre-school age to school age children year on year towards 2 to 16 year old coverage. In addition, a series of discrete geographical areas (pilot areas) offered LAIV vaccination to all primary school age children, allowing for a unique assessment and comparison of infection rates between pilot and non-pilot areas during roll-out.

Overall reductions in incidence rates of GAS and scarlet fever were observed within most of post-LAIV programme seasons when assessing the impact of the LAIV programme among the targeted (2 to 4 years and 5 to 10 years) and non-targeted groups using incidence rate ratios (IRRs) from Poisson regressions.

We assessed the overall effect of the pilot programme between the pre-introduction (2010/11-2012/13 influenza seasons) and post-introduction (2013/14-2016/17 influenza) periods using negative binomial regression by comparing the pre-to -post programme changes in incidence between the pilot and non-pilot areas (rIRR = ratio of incidence rate ratios). This showed significant reductions among the 5 to 10 years (rIRR of 0.57 (95% CI: 0.45 to 0.71; p-value: <0.001)); the 2 to 4 years (rIRR of 0.62 (95% CI:0.43 to 0.90; p-value: 0.011)) and the 11 to 16 years (rIRR of 0.63 (95% CI: 0.43 to 0.90; p-value: 0.018)) for GAS infections. A non-significant reduction was also seen for iGAS in 2-4 year olds (rIRR of 0.58 (95% CI: 0.21 to 1.65; p-value=0.31)). No difference was seen for iGAS 5 to 10 year olds, or for scarlet fever in both age groups (rIRRs (95% CI) of 1.1 (0.34-3.6), 0.96 (0.66-1.39), 1.16 (0.75-1.81) for iGAS age 5 to 10, scarlet fever age 2 to 4 and 5 to 10, respectively).

Our findings are compatible with the paediatric LAIV programme reducing the incidence of GAS and iGAS infections among children and support attaining high uptake of childhood influenza vaccination.

## Introduction

Group A *Streptococcus* (GAS) bacterium causes a range of clinical presentations including pharyngitis, scarlet fever, as well as focal and non-focal invasive group A streptococcal (iGAS) infection and pneumonia. Some of the latter have been identified as secondary bacterial infections associated with influenza [1-6].

In England, reports of scarlet fever have been low since the 1960s however in the winter of 2013/14, a resurgence was first observed and continued into 2015/16 [7-8]. A number of countries in Asia similarly reported increases in scarlet fever [9-10]. Both scarlet fever and invasive iGAS infection are notifiable diseases in England. A pronounced increase in scarlet fever and iGAS infection notifications, particularly in children under 10 years of age, is currently being seen during 2022 [11].

The paediatric live attenuated influenza (LAIV) intra-nasal immunisation programme in England commenced in the 2013/14 influenza season with the aim of reducing the annual burden of influenza by vaccinating all children aged 2 to 16 years to directly protect children themselves and indirectly protect others [12]. Since 2013/14, the programme has been incrementally rolled out by pre-school and school year group annually at the national level [13-15]. Simultaneously a number of geographically discrete pilot areas also began to vaccinate all primary school and some secondary school aged children which provided a unique opportunity to measure the direct and indirect impacts of the programme on the population. Previous studies of LAIV pilot areas have demonstrated significant differences in the reductions of influenza amongst the targeted age groups themselves and indirect effects due to reduction of influenza transmission on other age groups in pilot areas compared to non-pilot areas [13-17].

This study aims to investigate the potential epidemiological impact of the paediatric LAIV programme on the incidence of GAS infections including scarlet fever and iGAS by comparing infection rates in the period pre- and post-programme introduction in the LAIV pilot and non-pilot areas using data from the 2010-2011 to 2016-2017 influenza seasons in England.

## Methods

This study was a public health evaluation with data processed for the monitoring of the LAIV immunisation programme under Regulation 3 of the Health Service (Control of Patient Information) Regulations 2002.

### Data sources

Data on invasive and non-invasive GAS infections were extracted from a live national laboratory reporting system, Second Generation Surveillance System (SGSS), maintained by UK Health Security Agency (UKHSA, previously known as Public Health England) which collates all laboratory notifications of infectious diseases across England [18]. Data on scarlet fever based on clinical symptoms were extracted from the statutory notifications of infectious diseases (NOIDs) database, which contains statutory notifications submitted by diagnosing clinicians to local public health officials [19].

Individual level data on laboratory-confirmed GAS infections (all sample types) were extracted for the seasonal influenza surveillance period of each year (week 40 (October) to week 20 (May)) between 2010 and 2017. Pre-programme seasons (prior to the introduction of the LAIV programme) were defined to be 2010/11; 2011/12 and 2012/13 seasons and post-programme seasons were defined to be 2013/14; 2014/15; 2015/16 and 2016/17 (Supplementary Figure 1).

Individual level data were also postcode-matched to pilot or non-pilot areas at Local Authority (LA) level.

A pilot area was defined as a discrete geographical LA in England which participated in the full implementation of the roll out of the LAIV programme among all primary school age children (4 to 10 years) in their area in a given season [13-15] (Supplementary Figure 1).

A non-pilot area was defined as any LA not participating as a pilot area in the LAIV pilot programme which followed the national incremental roll out of the LAIV programme [13-15].

Data were categorised into age groups with LAIV targeted age groups being 2 to 4 years (all areas) and 5 to 10 years (pilot areas only) to assess the direct effects of the programme. It is important to note that the 2 to 4 years were offered the LAIV vaccine in both pilot and non-pilot areas from the beginning of the programme. Non-targeted age groups were categorised as < 2 years; 11 to 16 years; 17 to 44 years; 45 to 64 years; 65+ years to assess the indirect impact of the programme in these groups

Additionally, for the LAIV targeted age groups, data on invasive GAS, scarlet fever and iGAS (by restricting to include sample types depicting iGAS) notifications were also analysed.

Population denominators were derived from the Office for National Statistics’ yearly population estimates for each influenza season for each LA to generate denominators for pilot and non-pilot areas and by age group [20].

### Statistical analyses

Cumulative incidence rates were calculated as the number of GAS, scarlet fever, iGAS infections detected in a defined age group and season divided by the total population within the defined age group and pilot or non-pilot area. Exact Poisson confidence intervals (CI) were calculated.

Two analyses were performed. Firstly, incidence rate ratios (IRR) for GAS, scarlet fever or iGAS comparing pilot and non-pilot areas were estimated using Poisson regression within each season by age group.

Secondly, to assess whether the IRRs changed before and after programme implementation an indicator variable was created (0 for each of 2010/11 to 2012/13 and 1 for each of 2013/14 to 2016/17) and this was included in a negative binomial model along with pilot area as the interaction term. This gives a ratio of incidence rate ratios (rIRR = (post pilot incidence/pre pilot incidence)/ (post non-pilot incidence/ pre non-pilot incidence) where a value below 1 would indicate a potential impact of being in a pilot area on reducing rates. Negative binomial regression was used to allow for the fact that the incidence rate ratios varied within the pre and post implementation time periods.

All analyses were computed in STATA v15.

## Results

### LAIV targeted age groups (2 to 4 and 5 to 10 years)

In the pre-programme seasons, in both targeted age groups, incidence rates of all laboratory confirmed GAS infections were constantly greater in pilot areas than in non-pilot areas with IRRs of greater than 1.0 observed within each season (Figure 1A, Table 1A). Incidence rates of scarlet fever and iGAS in both targeted age groups in pilot and non-pilot areas in the pre-programme seasons varied and showed no significant differences with the exception of the 2012/13 season where rates were significantly lower in pilot areas for both age groups for scarlet fever (Figure 1B&C, Table 1B).

**TABLE 1:** Counts, crude incidence rates per 100,000 population (95% CI) and incidence rate ratios (IRR) (95% CI) of (A) all GAS, (B) scarlet fever and (C) invasive GAS infections by LAIV pilot and non-pilot areas and influenza seasons, 2010/11 to 2016/17 for targeted age groups, 2 to 4 years and 5 to 10 years *Note: significant IRRs with p-values <0.05 are highlighted in bold*

| Season/Pilot type | 2 to 4 years |  |  | 5 to 10 years |  |  |
| --- | --- | --- | --- | --- | --- | --- |
|  | Count | Crude rate per 100,000 population | Incidence Rate Ratio (95% CI) | Count | Crude rate per 100,000 population (95% CI) | Incidence Rate Ratio (95% CI) |
| 2010/11 |  |  |  |  |  |  |
| Pilot | 72 | 53.0 (41.5 to 66.8) | 1.44 (1.13 to 1.84) | 96 | 38.0 (30.8 to 46.4) | 1.59 (1.29 to 1.96) |
| Non-pilot | 667 | 36.8 (34.1 to 39.7) |  | 784 | 23.9 (22.3 to 25.7) |  |
| 2011/12 |  |  |  |  |  |  |
| Pilot | 78 | 56.7 (44.8 to 70.8) | 1.48 (1.17 to 1.87) | 119 | 46.7 (38.7 to 55.9) | 1.89 (1.56 to 2.30) |
| Non-pilot | 707 | 38.4 (35.6 to 41.3) |  | 819 | 24.7 (23 to 26.4) |  |
| 2012/13 |  |  |  |  |  |  |
| Pilot | 91 | 65.3 (52.6 to 80.2) | 1.05 (0.85 to 1.29) | 108 | 41.6 (34.2 to 50.3) | 1.24 (1.01 to 1.50) |
| Non-pilot | 1170 | 62.5 (58.9 to 66.1) |  | 1144 | 33.7 (31.8 to 35.7) |  |
| 2013/14 |  |  |  |  |  |  |
| Pilot | 107 | 75.9 (62.2 to 91.8) | 0.97 (0.80 to 1.18) | 146 | 54.7 (46.1 to 64.3) | 0.98 (0.83 to 1.16) |
| Non-pilot | 1481 | 78.1 (74.2 to 82.2) |  | 1952 | 55.7 (53.2 to 58.2) |  |
| 2014/15 |  |  |  |  |  |  |
| Pilot | 91 | 63.1 (50.8 to 77.5) | **0.65 (0.53 to 0.80)** | 136 | 49.5 (41.5 to 58.5) | 0.87 (0.73 to 1.04) |
| Non-pilot | 1885 | 97.2 (92.8 to 101.7) |  | 2043 | 56.7 (54.3 to 59.2) |  |
| 2015/16 |  |  |  |  |  |  |
| Pilot | 133 | 91.1 (76.3 to 108.0) | 0.83 (0.70 to 1.00) | 145 | 51.7 (43.6 to 60.8) | **0.84 (0.71 to 0.99)** |
| Non-pilot | 2124 | 108.7 (104.1 to 113.4) |  | 2282 | 61.7 (59.2 to 64.3) |  |
| 2016/17 |  |  |  |  |  |  |
| Pilot | 93 | 63.7 (51.4 to 78.0) | **0.72 (0.59 to 0.89)** | 130 | 45.2 (37.8 to 53.7) | **0.79 (0.67 to 0.95)** |
| Non-pilot | 1713 | 88.1 (84.0 to 92.4) |  | 2158 | 56.9 (54.5 to 59.4) |  |

(B) Scarlet fever
| Season/Pilot type | 2 to 4 years |  |  | 5 to 10 years |  |  |
| --- | --- | --- | --- | --- | --- | --- |
|  | Count | Crude rate per 100,000 population | Incidence Rate Ratio (95% CI) | Count | Crude rate per 100,000 population (95% CI) | Incidence Rate Ratio (95% CI) |
| 2010/11 |  |  |  |  |  |  |
| Pilot | 59 | 43.5 (33.1 to 56.1) | 0.85 (0.65 to 1.10) | 40 | 15.8 (11.3 to 21.6) | 0.73 (0.53 to 1.01) |
| Non-pilot | 931 | 51.4 (48.2 to 54.8) |  | 710 | 21.7 (20.1 to 23.3) |  |
| 2011/12 |  |  |  |  |  |  |
| Pilot | 89 | 64.7 (52.0 to 79.7) | 1.22 (0.98 to 1.52) | 50 | 19.7 (14.6 to 25.9) | 0.78 (0.59 to 1.04) |
| Non-pilot | 976 | 53.0 (49.7 to 56.4) |  | 833 | 25.1 (23.4 to 26.9) |  |
| 2012/13 |  |  |  |  |  |  |
| Pilot | 92 | 66.0 (53.2 to 81.0) | **0.78 (0.63 to 0.96)** | 76 | 29.3 (23.1 to 36.7) | **0.72 (0.57 to 0.90)** |
| Non-pilot | 1594 | 85.1 (81.0 to 89.4) |  | 1390 | 40.9 (38.8 to 43.2) |  |
| 2013/14 |  |  |  |  |  |  |
| Pilot | 305 | 216.5 (192.8 to 242.2) | 1.05 (0.93 to 1.18) | 290 | 108.6 (96.4 to 121.8) | 1.10 (0.98 to 1.24) |
| Non-pilot | 3922 | 206.92 (200.5 to 213.5) |  | 3447 | 98.3 (95.1 to 101.7) |  |
| 2014/15 |  |  |  |  |  |  |
| Pilot | 313 | 217.1 (193.7 to 242.5) | **0.74 (0.66 to 0.82)** | 232 | 84.4 (73.9 to 96.0) | **0.71 (0.62 to 0.81)** |
| Non-pilot | 5722 | 295.0 (287.4 to 302.7) |  | 4297 | 119.3 (115.7 to 122.9) |  |
| 2015/16 |  |  |  |  |  |  |
| Pilot | 364 | 249.3 (224.4 to 276.3) | **0.79 (0.71 to 0.87)** | 227 | 80.9 (70.7 to 92.1) | **0.60 (0.53 to 0.69)** |
| Non-pilot | 6206 | 317.5 (309.6 to 325.5) |  | 4980 | 134.7 (131.0 to 138.5) |  |
| 2016/17 |  |  |  |  |  |  |
| Pilot | 380 | 260.2 (234.7 to 287.7) | 1.04 (0.94 to 1.16) | 345 | 119.9 (107.6 to 133.3) | 1.15 (1.03 to 1.28) |
| Non-pilot | 4859 | 249.9 (243.0 to 257.1) |  | 3966 | 104.6 (101.4 to 107.9) |  |

(C) iGAS
| Season/Pilot type | 2 to 4 years |  |  | 5 to 10 years |  |  |
| --- | --- | --- | --- | --- | --- | --- |
|  | Count | Crude rate per 100,000 population | Incidence Rate Ratio (95% CI) | Count | Crude rate per 100,000 population (95% CI) | Incidence Rate Ratio (95% CI) |
| 2010/11 |  |  |  |  |  |  |
| Pilot | 1 | 0.7 (0.0 to 4.1) | 0.29 (0.04 to 2.10) | 3 | 1.2 (0.2 to 3.5) | 1.56 (0.47 to 5.16) |
| Non-pilot | 46 | 2.5 (1.9 to 3.4) |  | 25 | 0.8 (0.5 to 1.0) |  |
| 2011/12 |  |  |  |  |  |  |
| Pilot | 1 | 0.7 (0.0 to 4.1) | 0.45 (0.06 to 3.28) | 1 | 0.4 (0.0 to 2.2) | 0.59 (0.08 to 4.40) |
| Non-pilot | 30 | 1.6 (1.1 to 2.3) |  | 22 | 0.7 (0.4 to 1.0) |  |
| 2012/13 |  |  |  |  |  |  |
| Pilot | 6 | 4.3 (1.6 to 9.4) | 2.12 (0.90 to 5.02) | 1 | 0.4 (0 to 2.2) | 0.30 (0.04 to 2.16) |
| Non-pilot | 38 | 2.0 (1.4 to 2.8) |  | 44 | 1.3 (0.9 to 1.7) |  |
| 2013/14 |  |  |  |  |  |  |
| Pilot | 1 | 0.7 (0.0 to 4.0) | 0.36 (0.05 to 2.65) | 2 | 0.7 (0.1 to 2.7) | 0.82 (0.20 to 3.42) |
| Non-pilot | 37 | 2.0 (1.4 to 2.7) |  | 32 | 0.9 (0.6 to 1.3) |  |
| 2014/15 |  |  |  |  |  |  |
| Pilot | 0 | 0.0 (0.0 to 2.6) | - | 3 | 1.1 (0.2 to 3.2) | 0.91 (0.28 to 2.95) |
| Non-pilot | 54 | 2.8 (2.1 to 3.6) |  | 43 | 1.2 (0.9 to 1.6) |  |
| 2015/16 |  |  |  |  |  |  |
| Pilot | 5 | 3.4 (1.1 to 8.0) | 0.92 (0.37 to 2.27) | 2 | 0.7 (0.1 to 2.6) | 0.45 (0.11 to 1.83) |
| Non-pilot | 73 | 3.7 (2.9 to 4.7) |  | 59 | 1.6 (1.2 to 2.1) |  |
| 2016/17 |  |  |  |  |  |  |
| Pilot | 3 | 2.1 (0.4 to 6.0) | 0.71 (0.22 to 2.28) | 3 | 1.0 (0.2 to 3.1) | 1.24 (0.38 to 4.04) |
| Non-pilot | 56 | 2.9 (2.2 to 3.7) |  | 32 | 0.8 (0.6 to 1.2) |  |

**Figure 1.**
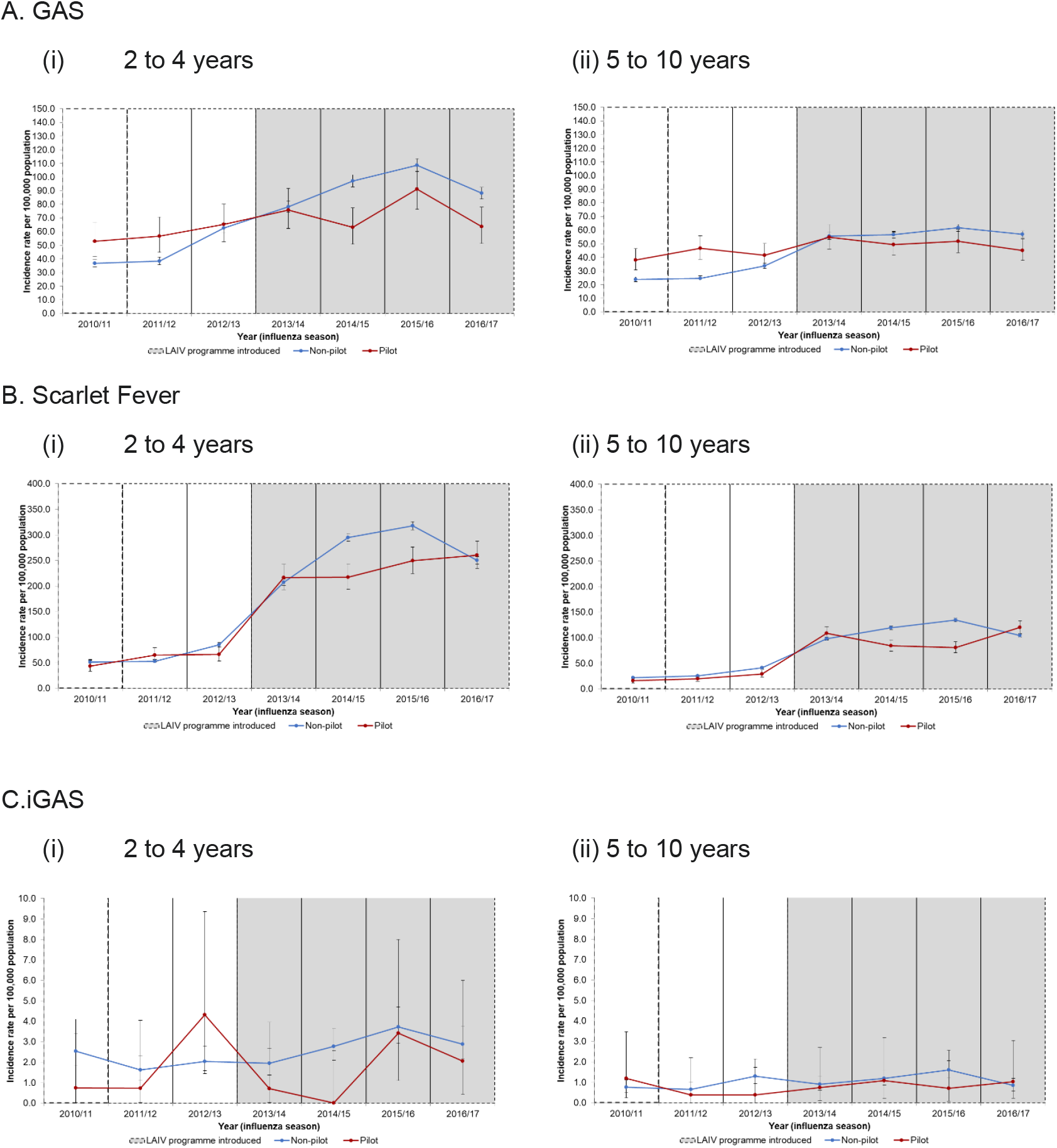
Incidence rates per 100,000 population (95% CI) of (A) GAS, (B) scarlet fever and (C) iGAS infections by LAIV pilot and non-pilot areas and influenza seasons (2010/11 to 2016/17) for targeted age groups (i) 2 to 4 years and (ii) 5 to 10 years

In the post-programme seasons, incidence rates of GAS, scarlet fever and iGAS in the 2 to 4 years and the 5 to 10 years were lower in pilot areas than in non-pilot areas with IRRs less than 1.0 observed within each season; with the exception of the 2013/14 and 2016/17 season for scarlet fever which saw slightly greater IRRs of 1.05 and 1.10 in the 2 to 4 years and 1.04 and 1.15 in the 5 to 10 years respectively; and in the 2016/17 season for iGAS with an IRR of 1.24 in the 5 to 10 years (Figure 1A(i)-C(i), Table 1A-C).

Significant reductions in incidence rates of GAS were noted in the 2 to 4 years in two of four post-programme seasons, 2014/15 and 2016/17 (IRR of 0.65 (95% CI: 0.53 to 0.80) and IRR of 0.72 (95% CI: 0.52 to 0.89) as well as in the 2015/16 and 2016/17 seasons for the 5 to 10 years (IRR of 0.84 (95% CI: 0.71 to 0.99) and IRR of 0.79 (95% CI: 0.67 to 0.95) (Table 1A). Significant reductions in incidence rates of scarlet fever in pilot areas were also noted among the targeted age groups, 2 to 4 years and 5 to 10, in post-programme seasons, 2014/15 and 2015/16 (2 to 4 years with IRRs of 0.74 (95% CI: 0.66 to 0.82) and of 0.79 (95% CI: 0.71 to 0.87); 5 to 10 years with IRRs of 0.71 (95% CI: 0.62 to 0.81) and of 0.60 (95% CI: 0.53 to 0.69) (Table 1C).

Comparing the changes pre-to post-programme in pilot and non-pilot areas for GAS using negative binomial regression showed that for 2 to 4 year olds the 1.26-fold increase in pilot areas was lower than the 2.03-fold increase in non-pilot areas (rIRR 0.62 (95% CI: 0.43 to 0.90; p-value 0.011)). In 5 to 10 year olds the 1.19 fold increase in pilot areas was lower than the 2.10 fold increase in non-pilot areas (rIRR 0.57 (95% CI: 0.45 to 0.71; p-value <0.001)). For iGAS the fold change was also lower in pilot areas for 2 to 4 year olds, but not significantly so (rIRR 0.58 (95% CI: 0.21 to 1.65; p-value=0.31)). For iGAS 5 to 10 year olds, and for scarlet fever in both age groups changes were similar in pilot and non-pilot areas (rIRRs (95% CI) of 1.1 (0.34-3.6), 0.96 (0.66-1.39), 1.16 (0.75-1.81) for iGAS age 5 to 10, scarlet fever age 2 to 4 and 5 to 10, respectively).

### Non-targeted age groups

Differences in incidence rates of GAS between pilot and non-pilot areas in the pre-programme seasons among the non-targeted groups were minimal and varied (Figure 2, Supplementary Table 1).

**Figure 2.**
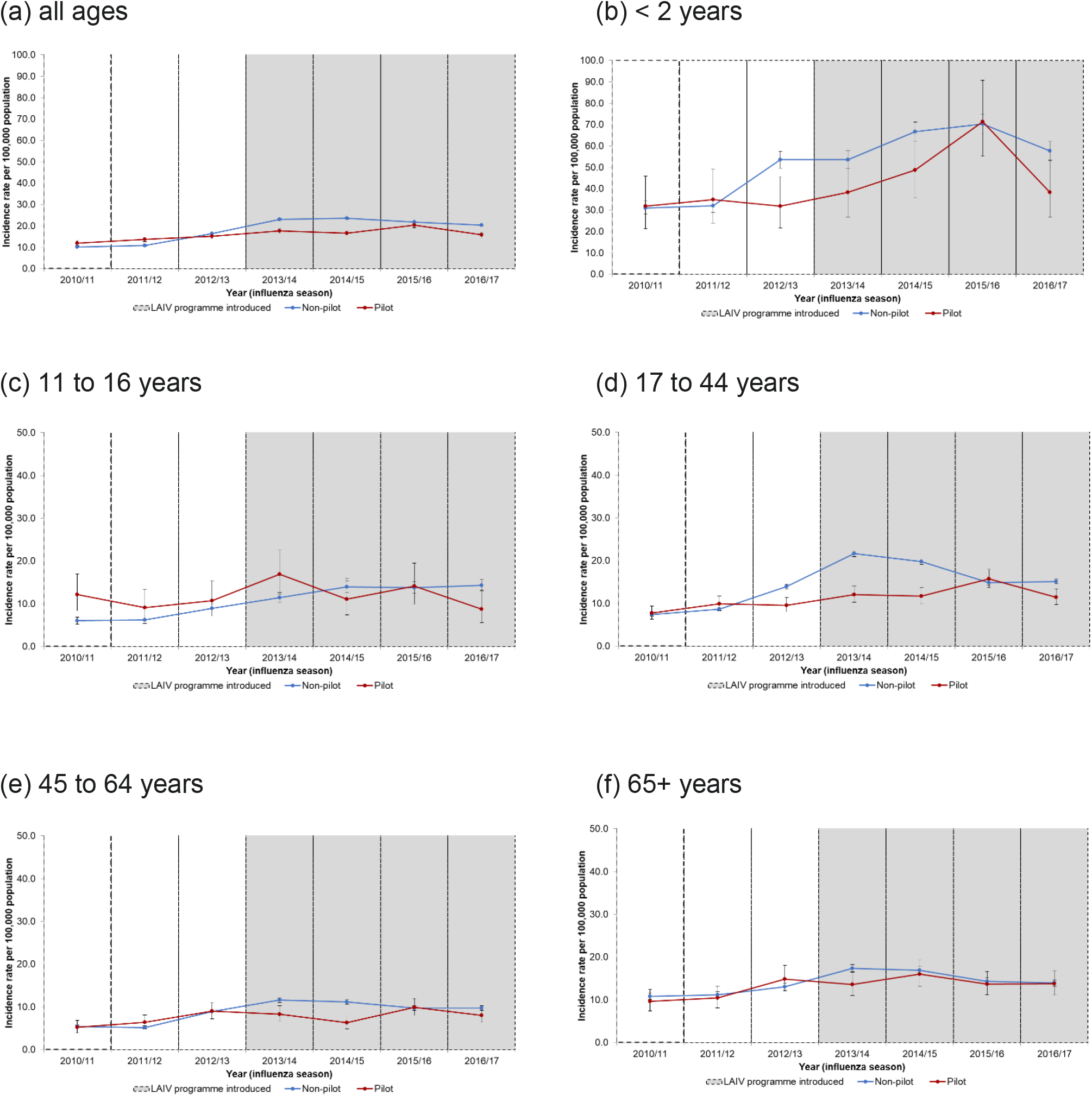
Incidence rates per 100,000 population (95% CI) of GAS infections by LAIV pilot and non-pilot areas and influenza seasons (2010/11 to 2016/17) for non-targeted age group (a) all ages (b) < 2 years (c) 11 to 16 years (d) 17 to 44 years (e) 45 to 64 years and (f) 65+ years

All non-targeted age groups, with the exception of all-ages and 65+ years, saw greater incidence rates of GAS among pilot areas in at least one of the post-programme seasons and IRRs greater than 1.0 (Figure 2, Supplementary Table 1). The all-age and 65+ years age groups observed lower albeit non-significant IRRs for GAS in pilot areas compared to non-pilot areas within all post-programme seasons (Supplementary Table 1).

Comparing the changes pre-to post-programme in pilot and non-pilot areas for GAS using negative binomial regression showed no significant differences except for age 11 to 16 where the 1.20 fold increase in pilot areas was lower than the 1.89 fold increase in non-pilot areas (rIRR 0.63 (95% CI: 0.43 to 0.92; p-value 0.018)). In the other ages increases were also lower in the pilot areas, but not significant (range of rIRRs from 0.73 to 0.94).

## Discussion

Our findings suggest that there were relative reductions in the incidence rates of group A streptococcal (GAS) for 2 to 4, 5 to 10 and 11 to 16 year olds in areas where the LAIV programme was piloted for these age groups compared to those that introduced the programme gradually. Reductions were also observed in incidence rates of GAS between pilot and non-pilot areas within seasons, in at least two of the post-programme influenza seasons in all age groups; with significant reductions noted in the 2 to 4 years and <2 years. Reductions were also noted when assessing scarlet fever and iGAS infection rates within seasons in the targeted age groups (2 to 10 years), but significant reductions were noted only for scarlet fever in both 2 to 4 and 5 to 10 years. These findings are consistent with a positive impact of the LAIV programme on reducing GAS infections.

Many prior studies have associated influenza to predispose to secondary bacterial infections particularly when influenza activity is high [2-6]. A review study summarised that increases in GAS and iGAS infections were noted in or after the 2009 influenza A(H1N1) pdm09 pandemic, and another study in England found that high influenza activity in the 2010/11 season contributed to an increased risk of concurrent invasive bacterial infections [2,6]. Prior findings from LAIV impact studies in England have shown a reduction in influenza infections among LAIV targeted age groups when comparing rates in pilot and non-pilot areas in England [13-16]. Our findings suggest that the reductions in influenza among children also contributes to a reduction in secondary bacterial infections.

The reduction in GAS infections was most obvious in the 5 to 10 years age groups, this is expected given that it is these age groups where vaccine coverage differed most between pilot and non-pilot areas (pilot areas having offered vaccination to all these age cohorts, non-pilot areas having offered vaccination sequentially each year to an additional year group) [13,15, 22]. While both pilot and non-pilot areas offered vaccination to 2 to 4 year olds, the finding of significant reductions in the rates of GAS in the 2 to 4 years post-introduction of the LAIV programme may be due to the higher LAIV vaccine uptake in these age groups in pilot areas in comparison to the uptake in non-pilot areas, as well as the indirect effect of vaccinating 5 to 10 year olds [22]. Both findings provide encouraging evidence that an increase in the LAIV vaccine uptake may reduce the incidence of GAS infections.

Indirect impacts were also noted among non-targeted age groups albeit not reaching statistical significance (other than in the age group most proximal to the pilot intervention ages) similar to previous impact studies [13-15]. The observation of higher infections in the 2015/16 season coincides with an increase in iGAS notifications in England that year and the predominant circulation of influenza A(H1N1)pdm09 which is known to mainly affect children, however, moderate influenza vaccine effectiveness of almost 60% was observed for LAIV that season among the 2 to 17 year olds and therefore the increase in iGAS notifications in that year may not only be attributable to influenza related factors [21-22]

There are key strengths to our study including the unique opportunity to compare the impact of the LAIV programme roll out due to its pilot programme and the use of national infection data. There are, however, a number of limitations to this study. As a population level ecological study, our results should be interpreted with caution as these cannot fully be attributed to a causal association to the LAIV vaccine only as there may be other contributing factors such as changes in laboratory testing and surveillance reporting over time, however we have mitigated this limitation by using both historical (pre- and post-LAIV programme data) and geographical controls (pilot and non-pilot areas). Our findings of the reductions in the LAIV targeted age groups further mitigates these limitations. Secondly, it is important to note that most GAS infections are not laboratory confirmed and there may be regional differences in the acquisition of swabs and reporting over time this and other “surveillance artefacts” could vary between pilot and non-pilot areas over time and could contribute to apparent differences in rates of GAS over time. Thirdly it is important to note that some LAIV pilot areas targeted vaccination among secondary school (children aged 11 to 16 years) children in the 2014/15 influenza season and this may have been reflected in our results. Lastly, both the national roll out and the pilot programme targeted 2 to 4 year olds from the commencement of the programme with pilot areas achieving higher LAIV vaccine uptake rates [22].

Our study suggests that vaccinating children with LAIV may reduce the incidence of GAS infections, including potentially life-threatening iGAS infections. In the context of the increasing incidence of GAS observed in England during the current 2022/23 season and the potential for similar increases elsewhere following the COVID-19 pandemic and easing of associated social distancing measures, our findings support maximising childhood influenza vaccine uptake.

## Data Availability

All data produced in the present work are contained in the manuscript. Data cannot be made publicly available for ethical and legal reasons, i.e. public availability would compromise patient confidentiality as data tables list single counts of individuals rather than aggregated data.

## Acknowledgements

The authors would like to thank Dr Richard G. Pebody for the original concept of this study and initiating this work.

## Author contributions

MS, FW, NA conceptualised the study and its design. Data collection and analysis were prepared by MS, FW, and NA. The first draft of the manuscript was written by MS. All authors commented, read and approved the final manuscript.

---

**Supplementary Figure 1.**
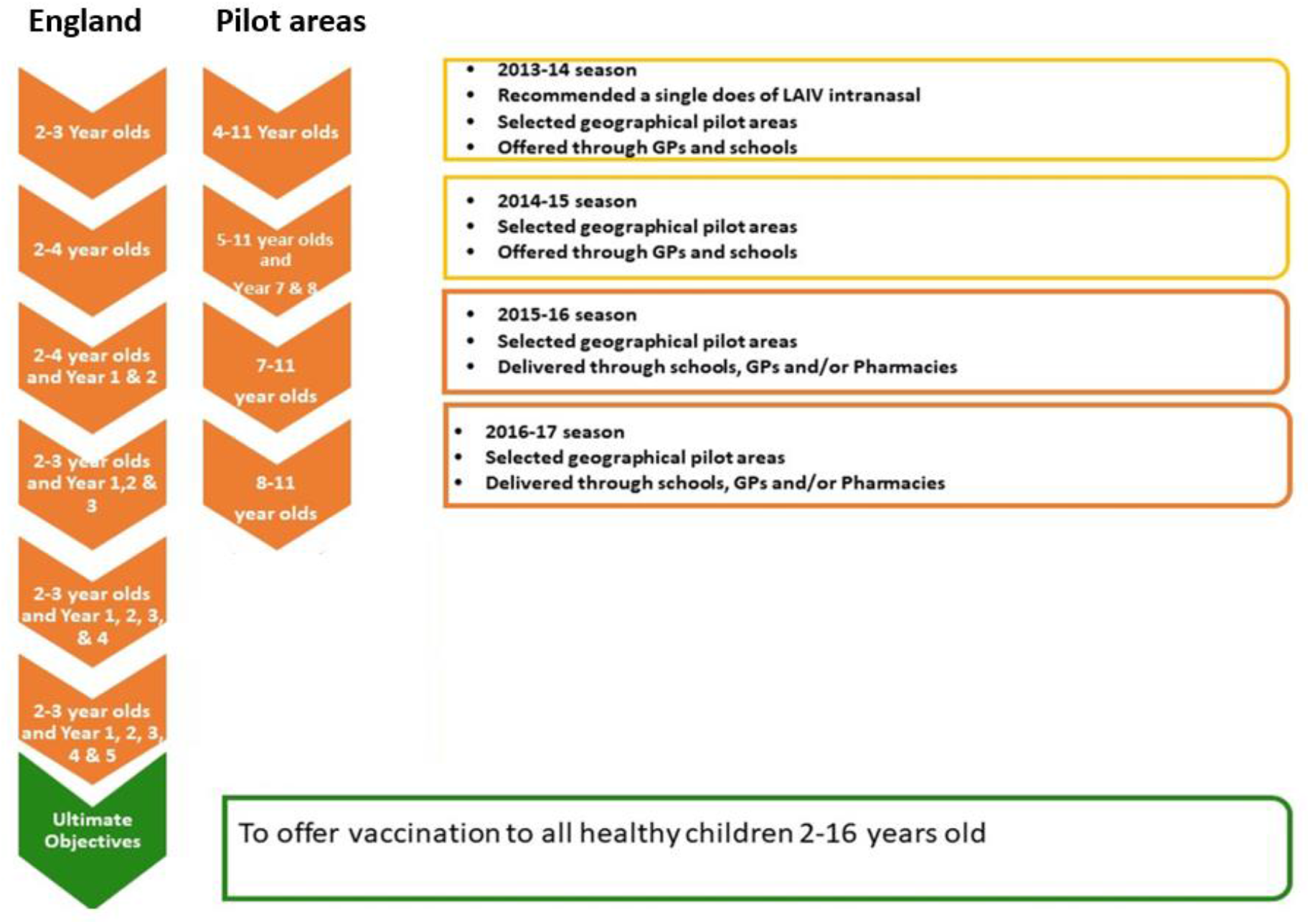
Illustrative figure of the roll out of LAIV programme, 2013/14 to 2016/17 seasons.

**Supplementary Table 1.** Counts, crude incidence rates per 100,000 population (95% CI) and incidence rate ratios (IRR) (95% CI) of GAS by LAIV pilot and non-pilot areas and influenza seasons, 2010/11 to 2016/17 for non-targeted age groups, (A) all ages, < 2 years and 11 to 16 years and (B) 17 to 44 years, 45 to 64 years and 65+ years *Note: significant IRRs with p-values <0.05 are highlighted in bold*

| Season/Pilot type | All ages |  |  | <2 years |  |  | 11 to 16 years |  |  |
| --- | --- | --- | --- | --- | --- | --- | --- | --- | --- |
|  | Count | Crude rate per 100,000 (95% CI) | Incidence Rate Ratio (95% CI) | Count | Crude rate per 100,000 (95% CI) | Incidence Rate Ratio (95% CI) | Count | Crude rate per 100,000 (95% CI) | Incidence Rate Ratio (95% CI) |
| 2010/11 |  |  |  |  |  |  |  |  |  |
| Pilot | 452 | 11.9 (10.9 to 13.2) | 1.18 (1.07 to 1.30) | 29 | 31.9 (21.3 to 45.8) | 1.03 (0.71 to 1.50) | 34 | 12.2 (8.4 to 17.0) | 2.01 (1.40 to 2.89) |
| Non-pilot | 4963 | 10.2 (9.9 to 10.4) |  | 385 | 31.0 (28.0 to 34.2) |  | 213 | 6.0 (5.3 to 6.9) |  |
| 2011/12 |  |  |  |  |  |  |  |  |  |
| Pilot | 523 | 13.7 (12.6 to 15.0) | 1.27 (1.17 to 1.39) | 32 | 34.9 (23.9 to 49.3) | 1.09 (0.76 to 1.56) | 25 | 9.1 (5.9 to 13.4) | 1.45 (0.96 to 2.19) |
| Non-pilot | 5332 | 10.8 (10.5 to 11.1) |  | 403 | 32.1 (29.0 to 35.3) |  | 219 | 6.3 (5.5 to 7.2) |  |
| 2012/13 |  |  |  |  |  |  |  |  |  |
| Pilot | 580 | 15.2 (13.9 to 16.4) | 0.93 (0.85 to 1.01) | 30 | 31.9 (21.6 to 45.6) | **0.60 (0.41 to 0.86)** | 29 | 10.7 (7.2 to 15.3) | 1.19 (0.81 to 1.74) |
| Non-pilot | 8148 | 16.4 (16.1 to 16.8) |  | 688 | 53.5 (49.6 to 57.6) |  | 310 | 9.0 (8.0 to 10.0) |  |
| 2013/14 |  |  |  |  |  |  |  |  |  |
| Pilot | 677 | 17.6 (16.3 to 19.0) | **0.77 (0.71 to 0.83)** | 36 | 38.4 (26.9 to 53.2) | 0.72 (0.51 to 1.00) | 45 | 16.9 (12.3 to 22.6) | 1.48 (1.09 to 2.01) |
| Non-pilot | 11516 | 23.0 (22.6 to 23.4) |  | 688 | 53.6 (49.7 to 57.7) |  | 389 | 11.4 (10.3 to 12.6) |  |
| 2014/15 |  |  |  |  |  |  |  |  |  |
| Pilot | 641 | 16.5 (15.2 to 17.8) | **0.70 (0.65 to 0.76)** | 45 | 48.8 (35.6 to 65.3) | **0.73 (0.54 to 0.99)** | 29 | 11.1 (7.4 to 15.9) | 0.79 (0.54 to 1.15) |
| Non-pilot | 11864 | 23.5 (23.1 to 24.0) |  | 835 | 66.6 (62.1 to 71.2) |  | 470 | 14.0 (12.7 to 15.3) |  |
| 2015/16 |  |  |  |  |  |  |  |  |  |
| Pilot | 798 | 20.4 (19.0 to 21.8) | 0.94 (0.87 to 1.01) | 66 | 71.3 (55.2 to 90.7) | 1.02 (0.79 to 1.31) | 37 | 14.1 (10.0 to 19.5) | 1.03 (0.73 to 1.44) |
| Non-pilot | 11072 | 21.8 (21.4 to 22.2) |  | 871 | 70.2 (65.6 to 75.0) |  | 463 | 13.8 (12.5 to 15.1) |  |
| 2016/17 |  |  |  |  |  |  |  |  |  |
| Pilot | 625 | 15.8 (14.6 to 17.1) | **0.78 (0.72 to 0.84)** | 36 | 38.3 (26.8 to 53.0) | **0.66 (0.47 to 0.93)** | 23 | 8.8 (5.6 to 13.2) | **0.61 (0.40 to 0.93)** |
| Non-pilot | 10458 | 20.4 (20.0 to 20.8) |  | 718 | 57.7 (53.5 to 62.1) |  | 484 | 14.3 (13.1 to 15.7) |  |

(B)
| Season/Pilot type | 17 to 44 years |  |  | 45 to 64 years |  |  | 65+ years |  |  |
| --- | --- | --- | --- | --- | --- | --- | --- | --- | --- |
|  | Count | Crude rate per 100,000 (95% CI) | Incidence Rate Ratio (95% CI) | Count | Crude rate per 100,000 (95% CI) | Incidence Rate Ratio (95% CI) | Count | Crude rate per 100,000 (95% CI) | Incidence Rate Ratio (95% CI) |
| 2010/11 |  |  |  |  |  |  |  |  |  |
| Pilot | 107 | 7.8 (6.4 to 9.4) | 1.05 (0.86 to 1.28) | 52 | 5.3 (3.9 to 6.9) | 0.97 (0.29 to 3.23) | 62 | 9.7 (7.4 to 12.4) | 0.89 (0.69 to 1.15) |
| Non-pilot | 1390 | 7.4 (7.0 to 7.8) |  | 665 | 5.4 (5.0 to 5.8) |  | 859 | 10.8 (10.1 to 11.6) |  |
| 2011/12 |  |  |  |  |  |  |  |  |  |
| Pilot | 137 | 9.9 (8.3 to 11.8) | 1.15 (0.96 to 1.36) | 64 | 6.4 (4.9 to 8.1) | 1.23 (0.95 to 1.59) | 68 | 10.4 (8.1 to 13.2) | 0.93 (0.73 to 1.20) |
| Non-pilot | 1634 | 8.7 (8.3 to 9.1) |  | 648 | 5.2 (4.8 to 5.6) |  | 902 | 11.2 (10.5 to 11.9) |  |
| 2012/13 |  |  |  |  |  |  |  |  |  |
| Pilot | 131 | 9.6 (8.0 to 11.3) | **0.68 (0.57 to 0.82)** | 90 | 9.0 (7.2 to 11.0) | 1.01 (0.81 to 1.25) | 101 | 14.9 (12.1 to 18.1) | 1.13 (0.93 to 1.39) |
| Non-pilot | 2621 | 14.0 (13.4 to 14.5) |  | 1118 | 8.9 (8.4 to 9.5) |  | 1097 | 13.1 (12.3 to 13.9) |  |
| 2013/14 |  |  |  |  |  |  |  |  |  |
| Pilot | 165 | 12.1 (10.3 to 14.1) | **0.56 (0.48 to 0.65)** | 83 | 8.3 (6.6 to 10.2) | **0.71 (0.57 to 0.89)** | 95 | 13.6 (11.0 to 16.6) | **0.78 (0.64 to 0.96)** |
| Non-pilot | 4053 | 21.6 (21.0 to 22.3) |  | 1458 | 11.6 (11.0 to 12.2) |  | 1495 | 17.4 (16.5 to 18.3) |  |
| 2014/15 |  |  |  |  |  |  |  |  |  |
| Pilot | 161 | 11.7 (10.0 to 13.7) | **0.59 (0.51 to 0.69)** | 64 | 6.3 (4.9 to 8.1) | **0.56 (0.44 to 0.73)** | 115 | 16.0 (13.3 to 19.3) | 0.95 (0.78 to 1.15) |
| Non-pilot | 3714 | 19.8 (19.2 to 20.5) |  | 1423 | 11.2 (10.6 to 11.8) |  | 1494 | 16.9 (16.1 to 17.8) |  |
| 2015/16 |  |  |  |  |  |  |  |  |  |
| Pilot | 216 | 15.7 (13.7 to 18.0) | 1.06 (0.92 to 1.21) | 101 | 9.9 (8.1 to 12.0) | 1.02 (0.83 to 1.24) | 100 | 13.7 (11.2 to 16.7) | 0.96 (0.78 to 1.17) |
| Non-pilot | 2795 | 14.9 (14.3 to 15.5) |  | 1252 | 9.7 (9.2 to 10.3) |  | 1285 | 14.3 (13.5 to 15.1) |  |
| 2016/17 |  |  |  |  |  |  |  |  |  |
| Pilot | 158 | 11.5 (9.7 to 13.4) | **0.76 (0.65 to 0.89)** | 83 | 8.0 (6.4 to 10.0) | 0.82 (0.66 to 1.02) | 102 | 13.8 (11.2 to 16.7) | 0.99 (0.81 to 1.21) |
| Non-pilot | 2843 | 15.1 (14.6 to 15.7) |  | 1270 | 9.8 (9.2 to 10.3) |  | 1272 | 13.9 (13.2 to 14.7) |  |

